# Portable Brain Computer Interface Sleep Monitor Compared with Polysomnography in Macroscopic and Microscopic Sleep Structures

**DOI:** 10.1101/2025.10.31.25339277

**Authors:** Zijian Wang, Chenhao Zhao, Xiaoyu Bao, Junbiao Zhu, Kaixiu Jin, Yue Wang, Xuehao Chen, Yuanqing Li

## Abstract

Accurate assessment of both macroscopic and microscopic sleep structure is critical for diagnosing sleep disorders and advancing personalized interventions. However, most existing portable sleep-monitoring devices, while validated at the macroscopic level, have not been benchmarked against PSG for microscopic sleep features. This study systematically evaluates a novel portable brain–computer interface (BCI) device, TH25, against PSG across multiple levels of sleep architecture, using an AASM-standard montage (F3, F4, E1, E2, A1, A2) with patch-based dry electrodes. Thirty-one adults underwent simultaneous overnight recordings using both PSG and TH25 systems. The system’s stable dry electrode design and full-night analysis—including natural artifacts—further highlight the high quality and reliability of the recorded signals under realistic conditions. At the macroscopic level, sleep staging performance of TH25 using an automatic sleep staging algorithm demonstrated high agreement with PSG (overall accuracy: 91.2%; Cohen’s kappa: 0.85). Key sleep metrics such as total sleep time, sleep efficiency, and stage distributions were closely aligned across devices. At the microscopic level, TH25 accurately detected sleep spindles and slow waves with comparable precision and F1-scores to PSG. These results demonstrate that the TH25 device provides a portable, cost-effective, and user-friendly solution for home-based sleep monitoring, while maintaining PSG-level accuracy.

## Introduction

Sleep is a fundamental physiological process that plays a vital role in cognitive, emotional, and physical health Benca [1996]. Accurate monitoring of sleep architecture is essential not only for diagnosing sleep disorders Berry et al. [2017] but also for understanding brain function and developing personalized interventions Fernandez and Lüthi [2020], Guillodo et al. [2020]. Polysomnography (PSG) remains the gold standard for sleep assessment, providing comprehensive recordings of EEG, EOG, EMG, respiratory, and cardiac activity Berry et al. [2017]. However, the high cost, technical complexity, and limited ecological validity of PSG restrict its use in large-scale screening, longitudinal monitoring, and home-based applications Kwon et al. [2021], Roomkham et al. [2018]. These limitations have spurred interest in developing portable, user-friendly alternatives for sleep assessment Younes et al. [2017], Zhang et al. [2023].

Sleep architecture encompasses both macro-architecture, defined as the organization of sleep stages (N1, N2, N3, REM) and their transitions across the night as assessed by PSG, and micro-architecture, referring to transient oscillatory events such as spindles and slow waves Djonlagic et al. [2021]. Microscopic events such as spindles and slow waves represent fundamental mechanisms of sleep stability and memory consolidation Fernandez and Lüthi [2020], Rasch and Born [2013], Klinzing et al. [2019], but their reliable detection requires high-quality EEG signals. However, many widely used commercial devices, including Dreem Arnal et al. [2020] and SleepProfiler Lucey et al. [2016], have primarily been validated at the macroscopic level, with little benchmarking against PSG for microstructural features. Therefore, validation studies should move beyond overall staging accuracy and explicitly assess a device’s ability to capture micro-level events, thereby establishing the fidelity of the recorded data for both research and clinical applicationsPurcell et al. [2017], Warby et al. [2014], Chinoy et al. [2021], Lee et al. [2019].

Recent advances in portable brain–computer interface (BCI) technology offer new opportunities for accessible, high-fidelity sleep monitoring Yoon and Choi [2023], Zhang et al. [2023]. Unlike conventional wearable trackers (e.g., wristbands or actigraphy) that rely mainly on actigraphy or peripheral signals, BCI devices directly record neurophysiological activity, enabling not only fine-grained analysis of both macro- and micro-architecture but also active closed-loop neuromodulation interventions Zhang et al. [2023], Fernandez and Lüthi [2020].

In this study, we evaluated the performance of a novel portable BCI device (TH25) compare with PSG. To demonstrate that the portable BCI device (TH25) can deliver high-quality signals comparable to PSG, we assessed both macrostructure (sleep staging) and microstructure (spindles, slow waves) through manual and algorithmic analyses. These findings highlight the potential of TH25 as a portable, cost-effective, and user-friendly solution for accurate home-based sleep monitoring and large-scale applications.

## Methods

### Participants

Thirty-one healthy adult volunteers (11 females and 20 males; mean age: 23.77 ± 3.51 years; age range: 18–33 years) were recruited through public advertisements and community outreach. Inclusion criteria included: age between 18 and 65 years, ability to use standard electronic devices, and voluntary participation with written informed consent after understanding the study procedures. Exclusion criteria were: presence of severe cardiopulmonary disease or psychiatric disorders (defined as a Hospital Anxiety and Depression Scale [HADS] score ≥ 11), known sleep disorders or neuromuscular conditions, recent use of sleep-related treatments (e.g., CPAP therapy, hypnotic medications, or surgery) within the past month, pregnancy or planned surgical intervention, or any other condition deemed inappropriate for participation by the investigators. The study protocol was approved by the Institutional Ethics Committee of South China Normal University, and written informed consent was obtained from all participants.

As shown in Table 1, participants exhibited mild anxiety (HAMA: 6.45 ± 5.67), moderate sleep disturbance (PSQI: 10.55 ± 5.05), and subthreshold insomnia symptoms (ISI: 5.77 ± 3.85). Elevated daytime sleepiness score was observed (ESS: 15.00±2.97), while depressive symptoms were minimal (BDI-Y: 5.65 ± 6.16).

**Table 1.** Demographics of the sample.

| | Mean ± SD | Min–Max |
| --- | --- | --- |
| Female/Male (n) | 11 / 20 |  |
| Age (years) | 23.77 ± 3.51 | 18–33 |
| BMI (kg·m<sup>-2</sup>) | 22.11 ± 2.13 | 17.72–26.12 |
| HAMA | 6.45 ± 5.67 | 0–23 |
| PSQI | 10.55 ± 5.05 | 1–21 |
| ISI | 5.77 ± 3.85 | 0–12 |
| ESS | 15.00 ± 2.97 | 9–21 |
| BDI-Y | 5.65 ± 6.16 | 0–19 |

*Note.* BMI, body mass index; HAMA, Hamilton Anxiety Scale; PSQI, Pittsburgh Sleep Quality Index; ISI, Insomnia Severity Questionnaire; ESS, Epworth Sleepiness Scale; BDI-Y, Beck Depression Inventory for Youth

### Study device

The TH25 is a lightweight, portable brain–computer interface (BCI) device designed for overnight sleep monitoring in home and clinical settings. The main processing unit (8 cm × 2 cm × 3 cm, 50g) is embedded in a flexible headband, ensuring user comfort throughout the night. It supports full-night EEG acquisition and simultaneously measures heart rate, oxygen saturation, and estimated respiratory rate using a MAX30102 sensor. The system integrates a 24-bit ADC with low noise (1*µV*) and high CMRR (100 dB), and communicates via Bluetooth for real-time data transmission. The device uses detachable Ag/AgCl gel electrodes with high biocompatibility and low contact impedance. The electrode–device connection is secured by precision snap-ons to ensure stable signal quality and ease of cleaning. For a detailed description of the hardware architecture, electrode design, and software features, see Supplementary Material 1.

### Experimental design

Participants first underwent a brief telephone screening followed by an in-person visit to the sleep laboratory, during which informed consent was obtained and eligibility was confirmed through a medical and lifestyle questionnaire. On the recording night, each participant completed an overnight study with simultaneous polysomnography (PSG) and the TH25 device, fitted by a registered technologist. The TH25 system employed patch-based dry electrodes in an AASM-standard montage (F3, F4, E1, E2, A1, A2) to ensure stable and high-fidelity recordings. Lights-off and lights-on times were self-selected by participants and used to define the recording window, which typically ranged from approximately 22:00 to 08:30. Importantly, full-night EEG was retained without discarding segments due to movement artifacts or transitional periods. PSG and TH25 streams were subsequently synchronized by resampling TH25 data onto PSG timestamps, allowing for epoch-by-epoch and event-level comparisons of both macroscopic sleep architecture and microscopic features such as spindles and slow waves.

### Polysomnographic assessment

Overnight Polysomnographic (PSG) recordings were conducted in a sound-attenuated, temperature-controlled sleep laboratory using a Grael 4k PSG–EEG Amplifier system (Compumedics, Australia), following the American Academy of Sleep Medicine (AASM) guidelines Berry et al. [2017]. The standard montage included electroencephalography (EEG: F3, F4, C3, C4, O1, O2, M1, M2), electrooculography (EOG), electromyography (EMG) of the submental and bilateral anterior tibialis muscles. 90 Sleep stages and respiratory events were manually scored by three registered sleep technologists according to the AASM criteria (version 2.6). All recordings were independently labeled by the three scorers. In cases of disagreement, the final annotation was determined through group discussion and consensus to establish the ground-truth reference standard.

In addition to standard sleep staging, sleep spindles and slow waves were automatically detected using the Profusion Sleep software (Compumedics, Australia). For spindle detection, the algorithm applied a bandpass filter of 11–16 Hz and a duration threshold of 0.5–2.0 seconds on central EEG channels (C3, C4). For slow-wave detection, a 0.5–2 Hz bandpass filter and amplitude threshold (*>* 75*µV*) were applied on frontal channels (F3, F4). All detected events were subsequently reviewed and manually corrected by the same three expert scorers to ensure annotation accuracy and consistency.

### Data analysis

#### Raw signal comparison

For raw EEG signal analysis, we extracted the F3–M2 channel from both PSG and TH25 recordings across the entire sleep period. First, the continuous EEG signals were segmented into consecutive 30-second epochs. Each epoch was labeled into one of the five standard sleep stages: Wake, N1, N2, N3, or REM. For each stage, representative raw waveforms and power spectral densities (PSDs) were derived from both systems to enable visual and spectral comparison of stage-specific EEG features. Power spectral density was estimated using Welch’s method, as defined in Equation (1):

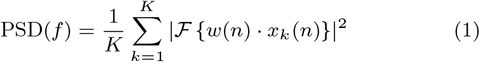

where *x*_*k*_(*n*) is the *k*-th windowed segment of the signal, *w*(*n*) is the window function, and ℱ denotes the Fourier transform. Second, we computed the relative power of four canonical EEG frequency bands—delta (0.5–4 Hz), theta (4–8 Hz), alpha (8–13 Hz), and beta (13–30 Hz)—across the entire night. The relative power for each band was calculated using Equation (2):

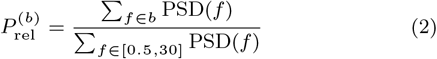

This yielded time-varying band power profiles for each device. To quantify the temporal agreement between PSG and TH25, we computed the mean percentage error (MPE) between their relative power trajectories over time, as shown in Equation (3):

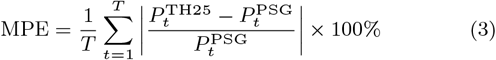

#### Macroscopic sleep parameters comparison

Macroscopic sleep architecture in this study was characterized based on the five-stage classification defined by the American Academy of Sleep Medicine (AASM) Berry et al. [2017], including Wake, N1, N2, N3, and REM sleep. To evaluate the sleep staging performance of the TH25 device, we conducted an epoch-by-epoch comparison with PSG-based ground truth using a confusion matrix. From this, standard classification metrics including precision, recall, F1-score, and Cohen’s kappa were computed for each sleep stage.

The macro-averaged F1-score across all classes was calculated using Equation (4):

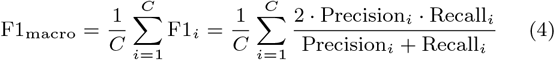

where *C* is the number of classes (i.e., five sleep stages), and F1_*i*_ is the F1-score for the *i*-th class.

Cohen’s kappa coefficient, which accounts for chance agreement, was calculated using Equation (5):

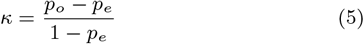

where *p*_*o*_ is the observed agreement and *p*_*e*_ is the expected agreement by chance.

Based on the full-night hypnograms from both PSG and TH25, a total of 20 macroscopic sleep parameters were extracted, including Sleep Period Time (SPT), Wake After Sleep Onset (WASO), and Total Sleep Time (TST), as summarized in Table 5. For each metric, we computed the mean and standard deviation across all participants for both systems. In addition, the absolute differences between TH25 and PSG values were calculated per subject, and their group-level mean and standard deviation were reported. These statistics were used to quantitatively assess the agreement and deviation between the two systems in capturing overall sleep architecture.

#### Microscopic sleep parameters comparsion

This study focused on two key microscopic sleep features: sleep spindles and slow waves. To evaluate the consistency between PSG and TH25, we computed the total number and density (events per minute) of spindles and slow waves detected by each device. Detection agreement was assessed using true positives (TP), false positives (FP), and false negatives (FN), with PSG annotations serving as the reference standard. A TH25-detected event was considered a true positive if it overlapped with a PSG-labeled event with an intersection-over-union (IoU) greater than 0.2, as defined in Equation (6):

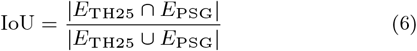

This metric ensures temporal alignment between events detected by the two systems, accounting for variations in event duration and onset. An illustration of the IoU-based event matching between TH25 and PSG for spindle detection is shown in Figure S6.

## Results

### Raw signals comparison

To evaluate signal-level consistency, the mean percentage error in relative spectral power across standard EEG frequency bands was computed between the TH25 and PSG (F3–M2), with PSG F3–M2 vs. F4–M1 used as a baseline comparison (Table2).

**Table 2.** Mean percentage error for *α, β, γ, andθ* EEG relative spectral power between the TH25 and PSG (F3–M2)

| Band | TH25/PSG(F3-M2) | PSG F3-M2/PSG F4-M1 |
| --- | --- | --- |
| Alpha (8–13 Hz) | 14.02 ± 9.24 | 18.72 ± 6.36 |
| Beta (13–30 Hz) | 23.72 ± 13.95 | 31.91 ± 18.41 |
| Delta (1–4 Hz) | 6.51 ± 5.05 | 8.27 ± 3.82 |
| Theta (4–8 Hz) | 13.23 ± 8.27 | 15.24 ± 3.76 |

Values are also provided for PSG1 (F3–M2) versus PSG2 (F4–M1) as a baseline.

The TH25 system showed lower or comparable spectral deviation from PSG than the inter-PSG (F3–M2 vs. F4–M1) benchmark. Specifically, the mean percentage error for alpha, beta, delta, and theta bands was 14.02% ± 9.24%, 23.72% ± 13.95%, 6.51% ± 5.05%, and 13.23% ± 8.27%, respectively—each lower than the corresponding inter-PSG differences (e.g., beta: 31.91% ± 18.41%).

In addition to quantitative spectral analysis, visual inspection of representative 30-second EEG epochs across all major sleep stages further illustrates the signal-level similarity between the portable BCI system and PSG (Fig. 1). The left panels show raw EEG waveforms recorded simultaneously from TH25 and PSG for Wake, N1, N2, N3, and REM stages. Despite differences in montage and amplitude scaling, both systems exhibit comparable morphological features characteristic of each sleep stage, including alpha attenuation during N1, sleep spindle and K-complex patterns in N2, high-amplitude slow waves in N3, and mixed-frequency activity in REM.

**Fig. 1:**
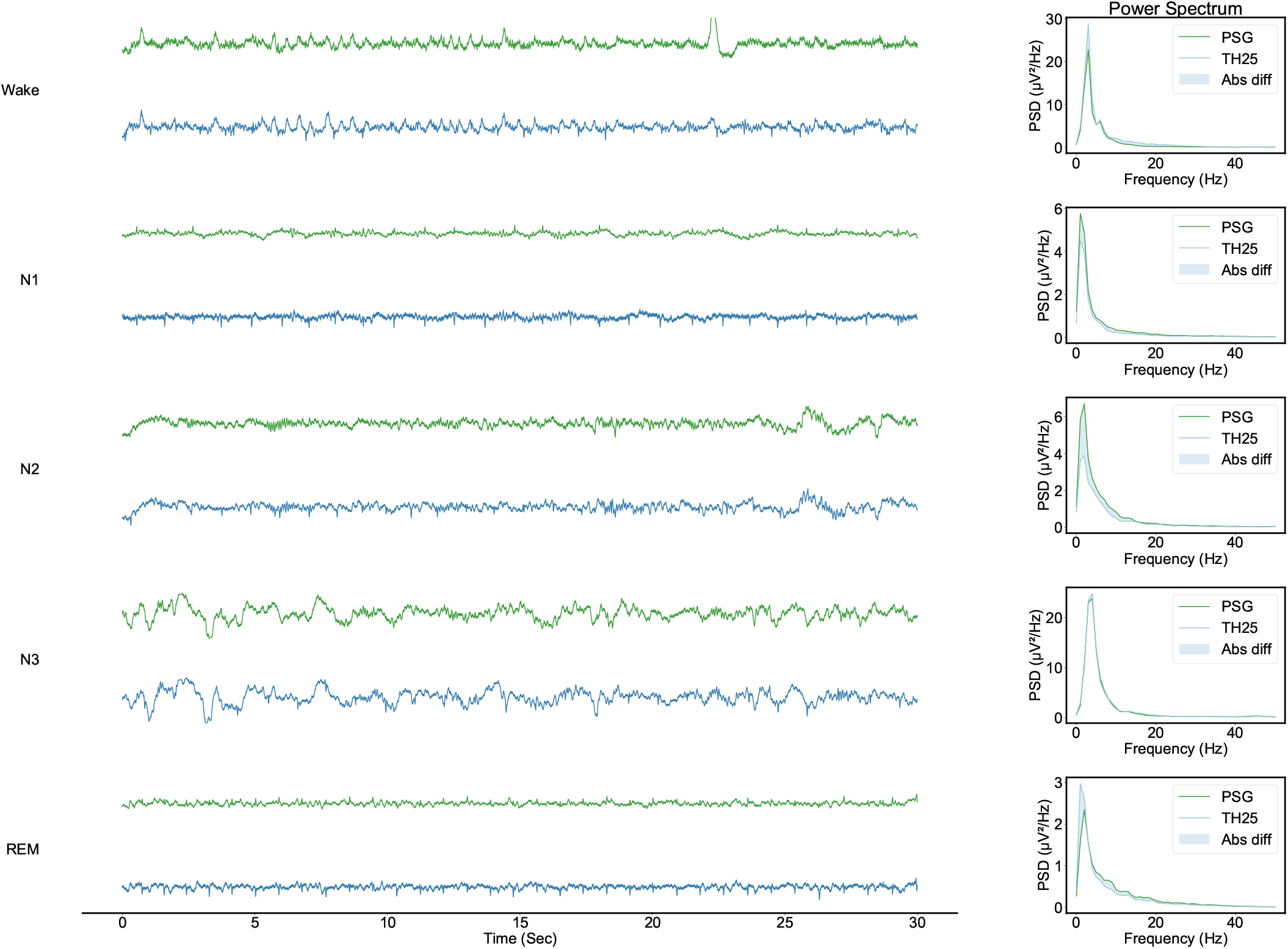
Representative 30-second EEG epochs and corresponding power spectral density (PSD) plots from five sleep stages (Wake, N1, N2, N3, REM), recorded simultaneously using the portable BCI system (TH25) and standard PSG (F3–M2). The signals are presented between −100 *µ*V and 100 *µ*V. For each stage, raw waveforms (left) show comparable morphology and oscillatory characteristics across systems. Power spectra (right) demonstrate consistent frequency-domain features, with stage-specific peaks preserved in both devices.

The corresponding power spectral density (PSD) plots on the right reveal consistent frequency profiles between TH25 and PSG across all stages. Peaks in the delta band during N3 and elevated theta–alpha activity in REM are preserved, indicating that TH25 reliably captures both time-domain and spectral characteristics of sleep EEG.

To further assess signal consistency over the entire night, we compared the relative spectral power time series between TH25 and PSG across five canonical frequency bands (delta, theta, alpha, beta, gamma). As shown in Fig. 2, the two systems exhibited similar temporal dynamics in all bands, with characteristic fluctuations corresponding to sleep stage transitions. Quantitatively, the mean percentage error (MPE) between TH25 and PSG was lowest in the delta band (10.35%), followed by theta (18.62%) and alpha (26.30%). Larger discrepancies were observed in the beta (61.22%) and gamma (45.44%) bands, likely reflecting higher sensitivity to muscle artifacts and hardware-specific differences in high-frequency response.

**Fig. 2:**
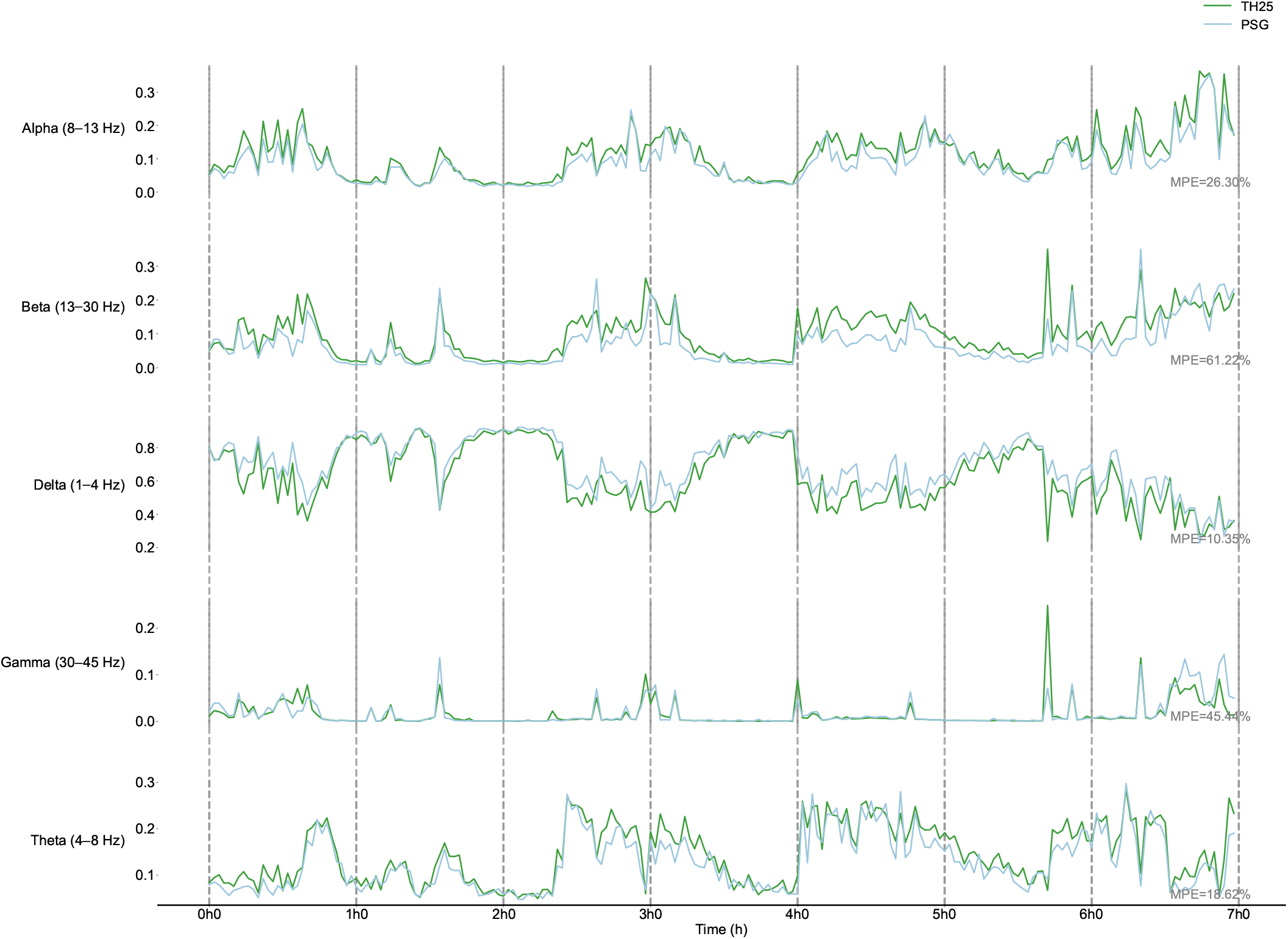
Overnight time series of relative spectral power across five canonical EEG frequency bands (delta: 1–4 Hz, theta: 4–8 Hz, alpha: 8–13 Hz, beta: 13–30 Hz, gamma: 30–45 Hz) recorded using the portable BCI system (TH25) and PSG. Each plot shows the temporal evolution of power in a given band over the course of the night. Mean percentage error (MPE) between TH25 and PSG is reported for each band, indicating close alignment in low-frequency bands (delta: 10.35%, theta: 18.62%) and larger deviations in higher-frequency bands (beta: 61.22%, gamma: 45.44%), likely due to inter-device differences in noise sensitivity and signal resolution. The results demonstrate that TH25 captures sleep-related spectral dynamics with high fidelity in physiologically relevant frequency ranges.

### Macroscopic sleep structure comparison

#### Agreement in sleep staging

Epoch-by-epoch analysis demonstrated substantial agreement between the portable TH25 system and PSG in sleep staging (Table 3, Fig. 3). The overall classification accuracy was 91.2% ± 5.0%, with a mean F1-score of 0.902 ± 0.073 and Cohen’s kappa of 0.847 ± 0.118. Stage-specific performance was highest in Wake (accuracy: 94.0% ± 5.0%; kappa: 0.891 ± 0.111) and N3 (accuracy: 90.8% ± 9.8%; kappa: 0.885 ± 0.109). Moderate agreement was observed in REM (accuracy: 83.4% ± 22.6%; kappa: 0.804 ± 0.221) and N2 (accuracy: 87.5% ± 14.5%; kappa: 0.819 ± 0.148). As expected, stage N1 showed the lowest performance, with an accuracy of 63.9% ± 23.8% and kappa of 0.626 ± 0.196.

**Table 3.** Agreement between PSG and portable BCI system in sleep staging (epoch-by-epoch analysis).

| Stage | Accuracy | Count | F1-score | Kappa |
| --- | --- | --- | --- | --- |
| ALL | 0.912 ± 0.050 | 1053 ± 165 | 0.902 ± 0.073 | 0.847 ± 0.118 |
| Wake | 0.940 ± 0.050 | 361 ± 219 | 0.933 ± 0.064 | 0.891 ± 0.111 |
| N1 | 0.639 ± 0.238 | 32 ± 20 | 0.636 ± 0.194 | 0.626 ± 0.196 |
| N2 | 0.875 ± 0.145 | 363 ± 140 | 0.874 ± 0.128 | 0.819 ± 0.148 |
| N3 | 0.908 ± 0.098 | 155 ± 56 | 0.898 ± 0.105 | 0.885 ± 0.109 |
| REM | 0.834 ± 0.226 | 140 ± 61 | 0.826 ± 0.203 | 0.804 ± 0.221 |

Accuracy and F1-score were computed for each sleep stage separately, based on epoch-by-epoch comparison between PSG and TH25. Count indicates the mean number of 30-second epochs labeled per stage.

**Fig. 3:**
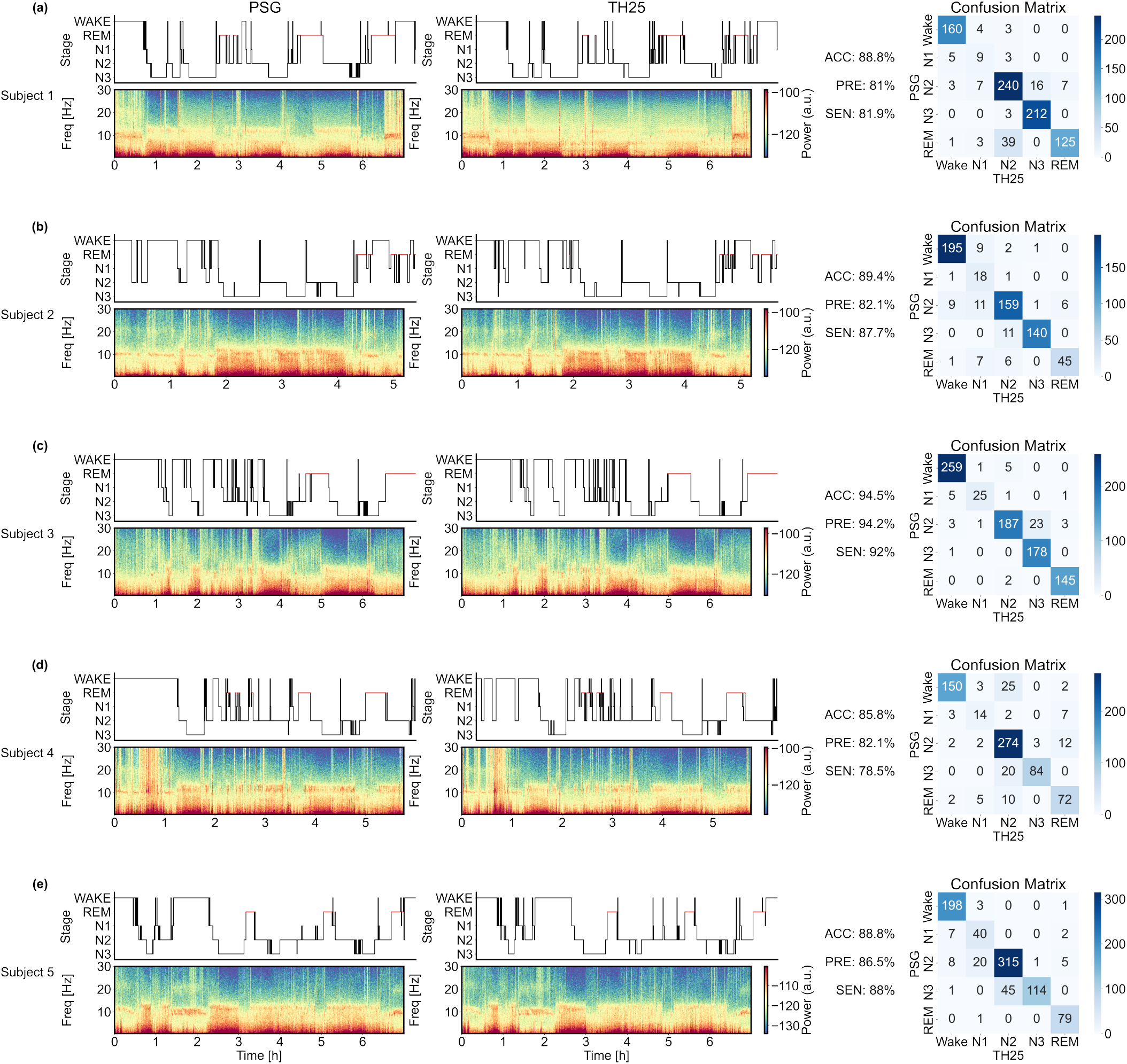
Representative sleep staging comparisons between PSG and the portable BCI system (TH25) across five participants. For each participant (row), the left and middle columns show overnight hypnograms and corresponding EEG spectrograms from PSG and TH25, respectively. The right column presents confusion matrices derived from epoch-by-epoch sleep stage classification, with PSG used as the reference.

Confusion matrices and representative hypnograms from five participants further illustrate the agreement between the portable BCI system and PSG (Fig.3). The top row in each case shows hypnograms generated by both systems, while the corresponding time–frequency plots of overnight EEG signals are displayed below. Right panels show confusion matrices, highlighting consistent staging performance across individuals.

Stage-wise durations were closely matched across systems, with mean differences of less than 2 minutes for N1, N2, N3, and REM sleep. Percentages of time spent in each stage (N1–REM) were similarly aligned, with differences typically below 1%. Latency metrics, including sleep onset latency (SOL) and REM latency, showed larger inter-individual variability but no systematic bias between systems.

### Microscopic sleep structure comparison

#### Agreement in sleep spindles and slow wave

The portable BCI system (TH25) demonstrated good performance in detecting sleep spindles and slow waves when compared to PSG (Table 4). For spindle detection, the mean sensitivity was ± 0.27, precision was 0.81 ± 0.26, and F1-score reached ± 0.26. For slow waves, detection metrics were similarly robust, with a sensitivity of 0.77 ± 0.19, precision of 0.78 ± 0.11, and F1-score of 0.77 ± 0.14. The mean number of spindles detected by TH25 (266.48 ± 233.09) was close to that of PSG (278.57 ± 256.71), with comparable event density per hour (30.46 ± 27.16 vs. 31.70 ± 29.57). Similarly, slow wave counts and densities were consistent between systems (TH25: 1449.86 ± 835.68, 162.99 ± 94.46/h; PSG: 1392.29 ± 752.38, 156.17 ± 84.35/h).These results suggest that TH25 is capable of reliably capturing key microstructural sleep features, showing near-equivalent event detection performance to standard PSG, particularly in the estimation of event density and temporal alignment within ±0.5 seconds.

**Table 4.** Detection performance of spindles and slow waves by the TH25 system compared to PSG.

| Metric | Spindle | SlowWave |
| --- | --- | --- |
| TP | 228 ± 225 | 1187 ± 779 |
| FP | 39 ± 73 | 263 ± 101 |
| FN | 51 ± 104 | 326 ± 260 |
| Sensitivity | 0.79 ± 0.27 | 0.77 ± 0.19 |
| Precision | 0.81 ± 0.26 | 0.78 ± 0.11 |
| F1-Score | 0.80 ± 0.26 | 0.77 ± 0.14 |
| Count (n, PSG) | 278.57 ± 256.71 | 1392.29 ± 752.38 |
| Count (n, TH25) | 266.48 ± 233.09 | 1449.86 ± 835.68 |
| Density (/h, PSG) | 31.70 ± 29.57 | 156.17 ± 84.35 |
| Density (/h, TH25) | 30.46 ± 27.16 | 162.99 ± 94.46 |

*Notes:* TP, FP, and FN refer to the mean and standard deviation of true positives, false positives, and false negatives, respectively, across all participants. Sensitivity, precision, and F1-score were calculated based on these counts using a temporal matching window of $\pm 0.5$ seconds between PSG-annotated and TH25-detected events. “Count” represents the number of events detected, and “Density” refers to the number of events per hour of total recording time.

**Table 5.** Comparison of sleep macrostructure parameters derived from the portable BCI system (TH25) and standard polysomnography (PSG).

|  | T25 | PSG | Difference |
| --- | --- | --- | --- |
| SPT(min) | 476.25 ± 71.03 | 465.89 ± 80.51 | 10.36 ± 107.37 |
| WASO(min) | 126.71 ± 86.27 | 119.82 ± 93.21 | 6.89 ± 127.01 |
| TST(min) | 349.54 ± 100.78 | 346.07 ± 103.47 | 3.46 ± 144.44 |
| Stage N1<br>duration(min) | 17.80 ± 9.37 | 16.30 ± 10.14 | 1.50 ± 13.81 |
| Stage N2<br>duration(min) | 183.54 ± 68.72 | 181.84 ± 70.17 | 1.70 ± 98.21 |
| Stage N3<br>duration(min) | 77.84 ± 31.06 | 77.68 ± 28.44 | 0.16 ± 42.12 |
| Stage REM<br>duration(min) | 70.36 ± 28.82 | 70.25 ± 30.97 | 0.11 ± 42.30 |
| Stage NREM<br>duration(min) | 279.18 ± 80.68 | 275.82 ± 84.15 | 3.36 ± 116.58 |
| SOL(min) | 41.98 ± 33.07 | 45.16 ± 38.47 | -3.18 ± 50.74 |
| Latency N1(min) | 81.54 ± 71.04 | 76.12 ± 63.40 | 5.41 ± 95.21 |
| Latency N2(min) | 46.62 ± 35.14 | 50.43 ± 42.78 | -3.80 ± 55.36 |
| Latency N3(min) | 69.68 ± 50.35 | 79.59 ± 55.79 | -9.91 ± 75.15 |
| Latency REM(min) | 186.00 ± 68.44 | 187.21 ± 62.37 | -1.21 ± 92.59 |
| Stage N1(%) | 5.12 ± 2.72 | 4.49 ± 2.29 | 0.64 ± 3.56 |
| Stage N2(%) | 51.80 ± 9.67 | 51.82 ± 9.42 | -0.02 ± 13.50 |
| Stage N3(%) | 23.14 ± 8.02 | 23.55 ± 7.70 | -0.41 ± 11.12 |
| Stage REM(%) | 19.93 ± 6.10 | 20.13 ± 6.93 | -0.20 ± 9.23 |
| Stage NEM(%) | 80.07 ± 6.10 | 79.87 ± 6.93 | 0.20 ± 9.23 |
| SE(%) | 66.77 ± 17.04 | 66.12 ± 17.77 | 0.65 ± 24.62 |
| SME(%) | 73.29 ± 17.42 | 74.73 ± 19.49 | -1.44 ± 26.14 |

Values represent the mean ± standard deviation across participants. Parameters include sleep period time (SPT), wake after sleep onset (WASO), total sleep time (TST), durations and proportions of each sleep stage (N1, N2, N3, REM), sleep onset latency (SOL), and latencies to each stage. SE = sleep efficiency; SME = sleep maintenance efficiency. NREM = N1 + N2 + N3. “Difference” refers to TH25 minus PSG.

## Discussion

Given the increasing need for reliable home-based sleep monitoring and intervention tools, portable BCI devices represent a significant advancement in sleep medicine Chaudhary et al. [2016], Al-Taleb et al. [2019], bridging clinical practice and personalized healthcare. Current portable sleep monitoring devices exhibit several notable limitations: (1) insufficient validation against PSG in microscopic sleep event detection, such as spindles and slow waves Edouard et al. [2021]. (2) instability of electrode attachment, often resulting in substantial signal dropout and excessive data rejection during overnight monitoring, as noted in prior validation studies(Arnal et al. [2020], Lucey et al. [2016]) (3) lack of validation for closed-loop neuromodulation, limiting their demonstrated utility to passive monitoring monitoring Ngo et al. [2013], Al-Taleb et al. [2019]. These limitations undermine data fidelity and limit their suitability for home-based, long-term applications.

This study systematically evaluated the performance of the portable brain–computer interface (BCI) device, TH25, against standard polysomnography (PSG), covering both macroscopic and microscopic aspects of sleep architecture. As PSG remains the clinical gold standard for sleep assessment, direct comparison is essential to establish the validity and reliability of alternative devices. Importantly, full-length overnight EEG recordings were analyzed without excluding segments affected by movement artifacts or transitional periods, thereby preserving the integrity of real-world sleep data.

From a macroscopic perspective, the TH25 system demonstrated high consistency with PSG across key sleep structure parameters (Table 5). Measures of total sleep time, sleep period time, wake after sleep onset, and sleep efficiency were closely aligned between the two systems, supporting the reliability of TH25 in assessing overall sleep quality and continuity. As summarized in Table 6, these results are comparable to—and in some respects exceed—the performance of other commercially available devices validated against PSG. For example, TH25 achieved higher staging agreement than the Dreem headband and outperformed both the Cognionics headband and SleepProfiler. This highlights its capacity to deliver PSG-comparable data quality for macroscopic sleep assessment in both research and clinical applications. Although the average sleep efficiency in this study appeared somewhat lower than expected, this was primarily due to the extended recording window (22:00–08:30) and the requirement to wear both PSG and TH25 devices, which prolonged quiet wakefulness. Nevertheless, participants still achieved physiologically adequate total sleep time, ensuring valid comparisons between systems.

**Table 6.**
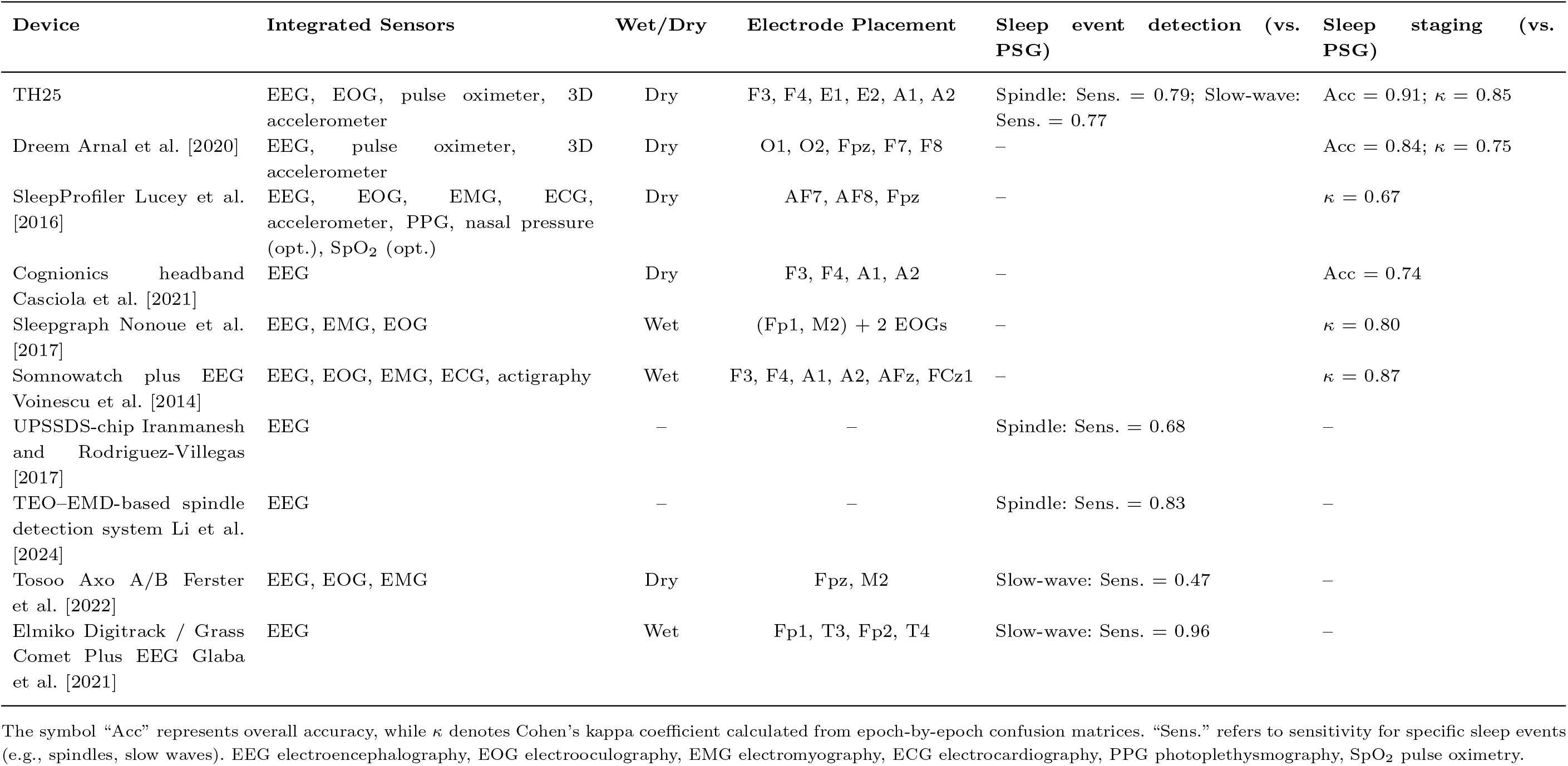
Comparison of wearable sleep monitoring devices with PSG.

At the microscopic level, the TH25 device also showed strong agreement with PSG in detecting sleep spindles and slow waves, which are critical biomarkers of sleep integrity and cognitive processing. Visual comparisons (Fig. 4) revealed that the morphology, temporal alignment, and oscillatory profiles of both event types were highly similar between systems across individuals. Quantitative results (Table 4) confirmed this consistency, with high sensitivity, precision, and F1-scores for both event types. Minor differences in spindle amplitude were noted, potentially due to differences in electrode montage and impedance, but the waveform structure and timing were well preserved. Notably, while several portable sleep-monitoring devices have incorporated spindle and slow-wave detection capabilities, most have not rigorously validated these microscopic features against PSG, with evaluations typically restricted to macroscopic sleep parameters or overall staging accuracy Sawangjai et al. [2019], de Gans et al. [2024], Arnal et al. [2020]. As shown in Table 6, the TH25 device is distinct in providing systematic validation at both the macroscopic and microscopic levels, achieving high agreement with PSG in sleep staging (accuracy = 0.91, *κ* = 0.85) as well as reliable detection of spindles and slow waves (sensitivity 0.79 and 0.77, respectively). Thus, this study addresses an important research gap by providing a direct and detailed validation of microscopic sleep features, reinforcing the TH25 system’s credibility for high-resolution, event-level sleep analysis.

**Fig. 4:**
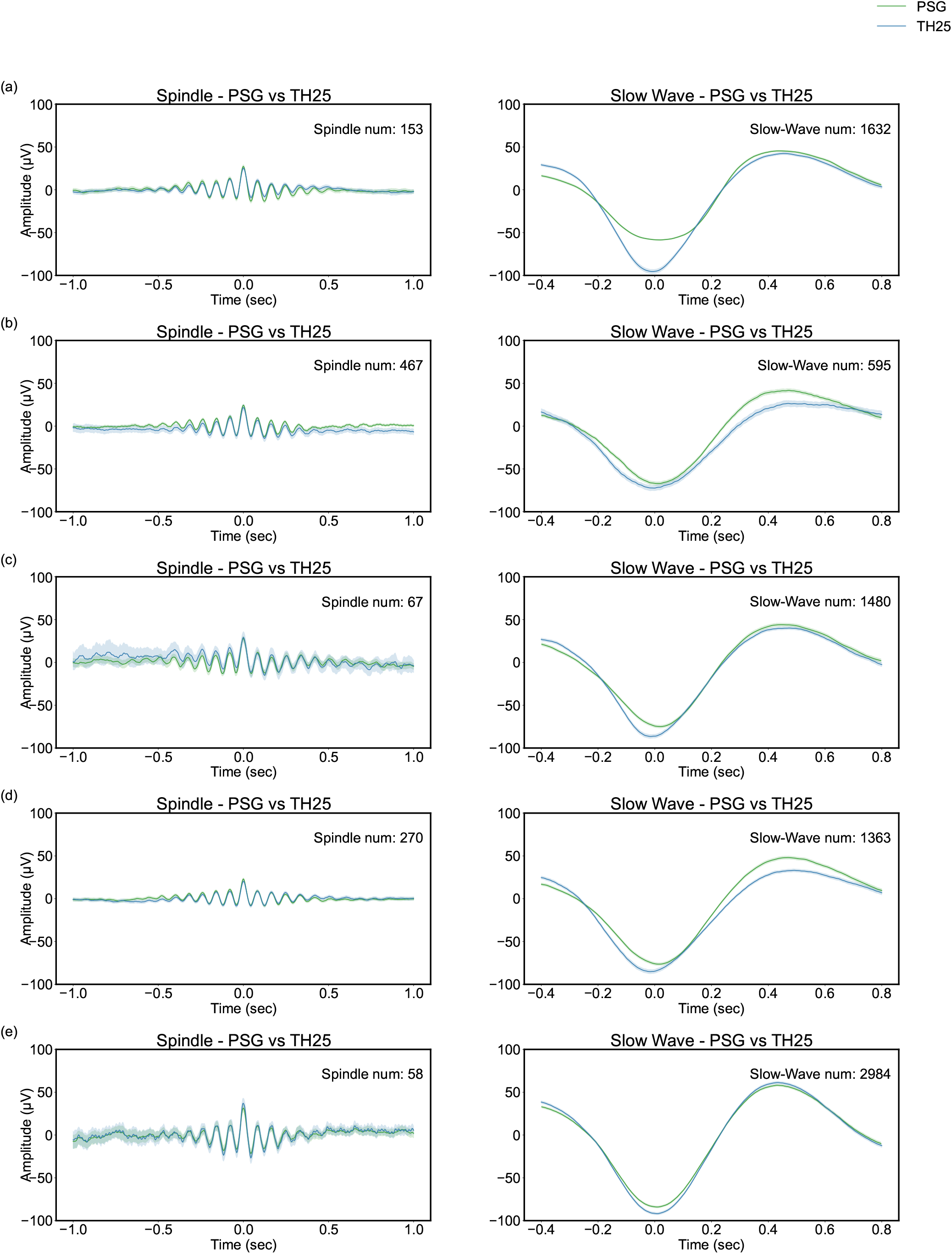
Comparison of spindle and slow-wave detection results between PSG and the TH25 system across five participants. Subplots (a)–(e) present the averaged waveforms over the whole night, with the number of events indicating the count of characteristic waves successfully matched between PSG and TH25 for each subject.

**Fig. 5:**
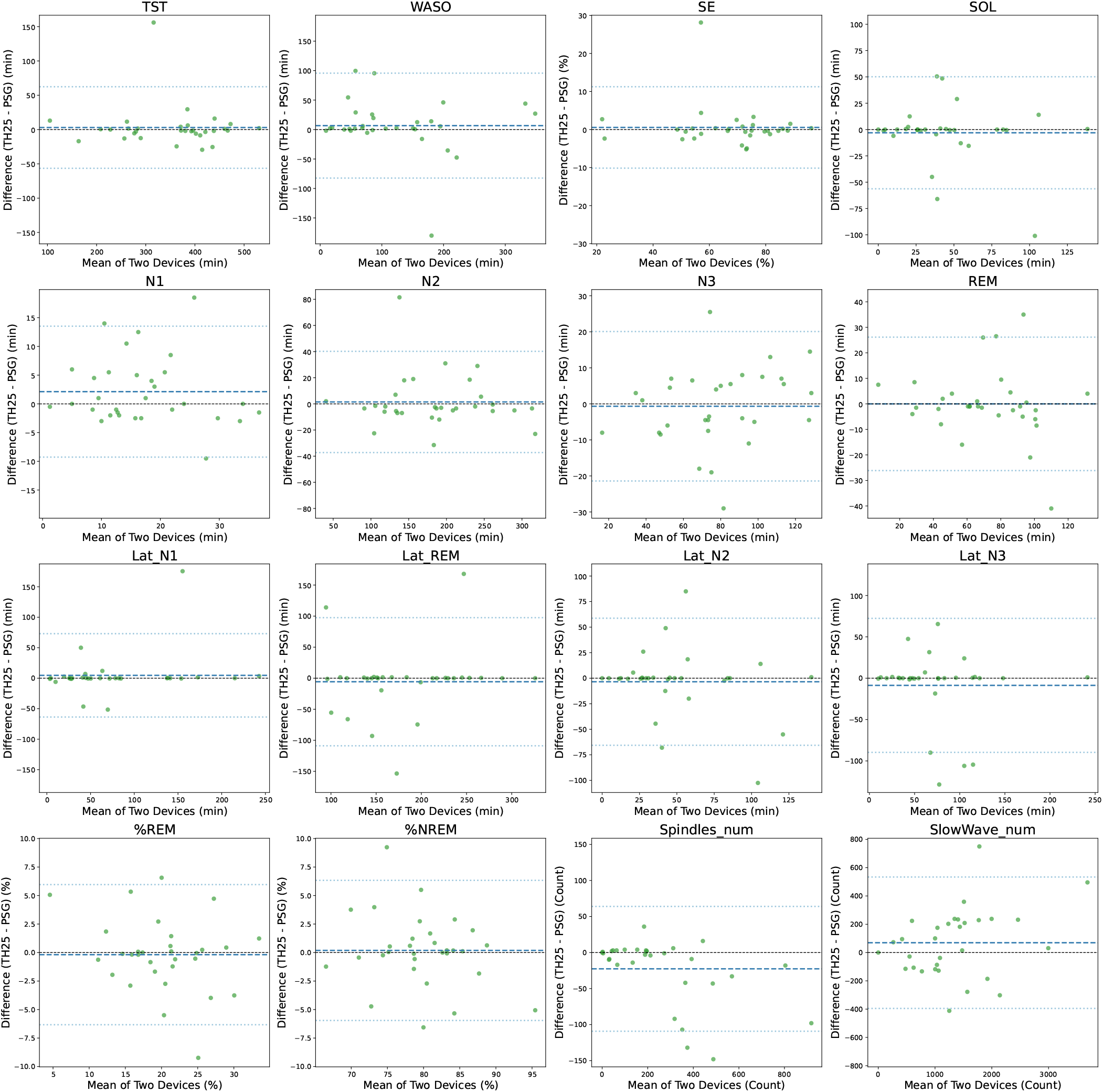
Bland–Altman plots comparing key macroscopic and microscopic sleep parameters between the TH25 and PSG systems. Each subplot shows the difference between TH25 and PSG against the mean of the two methods for a specific parameter. Parameters include TST, WASO, SE, SOL, stage durations (N1, N2, N3, REM), stage latencies (Lat N1, Lat N2, Lat N3, Lat REM), percentage of REM and NREM sleep, and the total number of spindles and slow waves. Solid lines indicate mean bias, and dashed lines indicate the 95% limits of agreement.

As summarized in Table 6, existing wearable devices differ substantially in electrode configuration and sensor integration, which directly constrains their ability to capture microstructural sleep events. For example, systems such as Dreem and SleepProfiler rely on limited frontal placements that are adequate for coarse staging but attenuate the morphology of spindles and slow waves. In contrast, the TH25 integrates patch-based dry electrodes with an AASM-standard montage (F3, F4, E1, E2, A1, A2), combining long-term stability, comfort, and standardized coverage. This hardware design not only minimizes signal dropout and electrode detachment but also ensures PSG-comparable fidelity at both macro- and micro-levels, thereby providing data quality suitable for research and clinical applications.

Importantly, beyond passive monitoring, the TH25 system also enabled reliable capture of neural responses to closed-loop auditory stimulation during sleep, further underscoring its utility as a Sleep intervention platform. This closed-loop feature was further examined in an additional experiment, in which auditory stimulation was delivered during stable N3 sleep while participants wore the TH25 device. Auditory stimulation delivered at the up-phase of slow oscillations elicited robust enhancements in slow-wave activity, with increased peak-to-peak amplitudes and elevated delta-band power relative to sham (Figure S9). These results are consistent with prior closed-loop stimulation studies Ngo et al. [2013], Papalambros et al. [2017], Henin et al. [2019], confirming that TH25 not only provides high-fidelity monitoring but also supports active neuromodulation paradigms with potential cognitive benefits.

A limitation of this study is that all participants were healthy adults with a relatively narrow age range(18-33years), which restricts the generalizability of the findings despite being comparable to previous validation studies Markov et al. [2024]. Future research should validate the TH25 device across larger and more diverse cohorts, including patients with obstructive sleep apnea (OSA) and other sleep disorders, to establish its broader clinical applicability. In addition, while this study demonstrated the feasibility of closed-loop neuromodulation using the BCI framework, further optimization of stimulation protocols and system integration will be necessary to fully realize the potential of portable BCI technologies in active sleep intervention.

This study underscores the robustness and practicality of the TH25 portable BCI device under realistic sleep conditions. The study retained full-length overnight EEG recordings without discarding any segments due to movement artifacts or transitional periods. Perturbations commonly encountered in real-world sleep experiments—such as the adaptation phase before sleep onset, gross body movements during sleep, and extended wake periods in bed after awakening—were all preserved to comprehensively assess the device’s performance in naturalistic settings. Additionally, the TH25 device employed patch-based dry electrodes(with AASM-standard montage, ensuring comfort, stability, and high-fidelity full-night recordings without electrode detachment or data loss.

## Conclusion

In this study, we conducted a comprehensive evaluation of the TH25 portable BCI device by benchmarking its performance against gold-standard polysomnography (PSG) at multiple levels. Specifically, we compared raw EEG signals, validated key macroscopic sleep architecture metrics such as total sleep time and sleep efficiency, and assessed microscopic features including spindle and slow wave detection.

In conclusion, this study provides systematic validations of a portable BCI device against PSG at both macroscopic and microscopic levels of sleep architecture, addressing a critical gap in prior wearable device research that has largely overlooked microstructural benchmarking. By demonstrating reliable detection of spindles and slow waves in addition to robust staging accuracy, the TH25 establishes that portable systems can achieve PSG-comparable fidelity. These findings highlight the potential of TH25 as a cost-effective, home-based, and high-precision brain–computer interface (BCI) device, providing scalable solutions for large-scale sleep monitoring and real-world clinical applications.

## Author contributions statement

Zijian Wang: Conceptualization, Methodology, Software, Validation, Writing – original draft. Chenhao Zhao: Data curation, Formal analysis, Visualization, Writing – review & editing. Xiaoyu Bao: Methodology, Investigation, Resources. Junbiao Zhu: Software, Validation, Data curation. Kaixiu Jin: Formal analysis, Visualization. Yue Wang: Resources, Project administration, Writing – review & editing. Xuehao Chen: Supervision, Validation, Writing – review & editing. Yuanqing Li: Conceptualization, Funding acquisition, Project administration, Supervision, Writing – review & editing.

## Acknowledgments

This work was supported in part by the STI 2030–Major Projects, China under Grant 2022ZD0208900; in part by the Key Research and Development Program of Guangdong Province, China under Grant 2018B030339001;

## Data Availability Statement

Data Availability Statement The datasets generated in this study, including the simultaneously recorded PSG data, TH25 portable BCI device data, and corresponding annotations, are available from the corresponding author upon reasonable request. Requests for access should be directed to Prof. Yuanqing Li.

