## Supplementary Information for "Portable Brain Computer Interface Sleep Monitor Compared with Polysomnography in Macroscopic and Microscopic Sleep Structures"

### Supplementary Material

#### Supplementary 1. Hardware

##### 1.1 TH25 Device

The TH25 is a portable brain-computer interface (BCI) device specifically designed for sleep monitoring applications. The physical appearance and main components of the device are illustrated in Figure S1, with detailed technical specifications provided in Table S1.

The device features a compact and ergonomic design with the main processing unit housed in a rigid black enclosure that measures approximately 8 cm × 2 cm × 3 cm and weighs 50 g. This lightweight form factor ensures comfortable overnight wear without exerting pressure on recruitments. The processing unit is seamlessly integrated into a flexible headband configuration, with the "HNNK" branding visible on the curved headband portion that contacts the subject's forehead. The device offers a battery life of at least 12 hours, sufficient for full overnight EEG monitoring.

The TH25 establishes wireless connectivity with smartphones, tablets, or other compatible devices through Bluetooth technology, enabling real-time data transmission. Under low electromagnetic interference conditions, the data packet loss rate is negligible. Key technical performance indicators include: average short-circuit noise level of 1 $\mu$V, 24-bit analog-to-digital converter (ADC) resolution, and common mode rejection ratio (CMRR) of 100 dB. The headband incorporates an integrated MAX30102 sensor operating at 50 Hz sampling rate, capable of directly measuring heart rate and blood oxygen saturation levels, while approximately inferring respiratory frequency from photoplethysmography (PPG) waveforms through embedded algorithms.

The robust design of the device minimizes motion artifacts and maintains signal integrity during natural sleep movements. Real-time data pre-processing includes digital filtering and artifact rejection, providing clean physiological signals for subsequent sleep structure analysis.

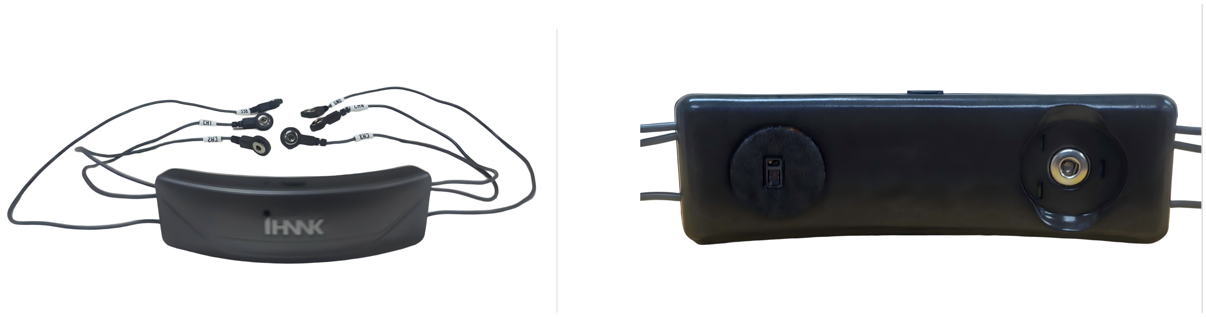

Figure S1: TH25 Brain-Computer Interface Device

**Table S1. TH25 Device Technical Specifications**

| **Parameter** | **Specification** |
|---|---|
| Dimensions | 8 cm $\times$ 2 cm $\times$ 3 cm |
| Weight | 50 g |
| Sampling rate | 250 Hz |
| Sampling resolution | 24 bit |
| Form Factor | Flexible headband with rigid processing unit |
| Battery Life | at least 12 hours (continuous operation) |
| Wireless Communication | Bluetooth |
| Data Transmission | Real-time streaming |
| ADC Resolution | 24-bit |

#### 1.2 Electrode System

The TH25 employs a six-electrode acquisition system designed for comprehensive sleep monitoring. Six independent wired connections extend from the main processing unit to specialized electrode connectors, each clearly labeled (CH1, CH2, CH3, CH4, GND, REF) and featuring a secure snap-on mechanism designed to interface with disposable gel electrodes. The electrode configuration follows the international 10-20 system placement: ground electrode (GND) connects to the right mastoid (M1), reference electrode (REF) connects to the left mastoid (M2), and the four EEG channels (CH1, CH2, CH3, CH4) are positioned at O2, Fp2, Fp1, and O1 locations respectively, covering frontal and occipital regions critical for sleep stage assessment.

The system utilizes disposable patch electrode featuring a dual-sided design optimized for bioelectric signal acquisition (Figure S3). The electrode's skin-contact surface (bottom) is covered with a conductive hydrogel layer that ensures stable electrical coupling with the skin while maintaining optimal impedance levels throughout extended recording periods. The top surface features a central metallic contact point surrounded by a non-conductive adhesive border, with the metallic contact designed to interface securely with the device's snap-on connectors.

Signal acquisition occurs through a multi-step process: prior to electrode placement, the subject's skin at designated locations is prepared with abrasive gel application to remove dead skin cells and reduce skin impedance, ensuring optimal signal quality. Following skin preparation, the electrodes are adhered to the subject's skin at the specified anatomical locations, establishing direct electrical contact through the conductive gel medium. The device's labeled electrode connectors (CH1-CH4, GND, REF) are then securely attached to the metallic contact points on the electrode surface, creating a complete electrical pathway from skin to analog front-end. This configuration enables high-fidelity bioelectric signal transmission with minimal contact impedance and artifact generation. Each EEG channel

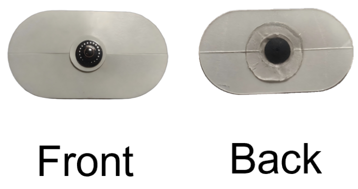


Figure S2: Disposable patch electrode

operates at 250 Hz sampling rate.

The electrode system is designed to maintain stable impedance levels throughout overnight recordings, with the conductive gel ensuring consistent signal quality even during subject movement. The snap-on connector design allows for quick and secure electrode attachment while minimizing connection artifacts that could compromise signal integrity.

### Supplementary 2. Software

#### 2.1 Sleep App

The Sleep App serves as the mobile interface for the TH25 system, supporting real-time sleep monitoring, automated sleep report visualization, in-app questionnaire completion (e.g., ISI, STAI, PedsQL), and secure data upload to the cloud. As shown in Figure S3, the app provides an intuitive interface for both passive tracking and active intervention. In addition to passive monitoring, it incorporates a closed-loop pink noise stimulation module that delivers phase-locked auditory cues during deep sleep to enhance slow-wave activity, as well as a music-assisted sleep function that generates personalized soundscapes to promote sleep onset and relaxation. Furthermore, the Sleep App is fully compatible with major mobile operating systems, including Android and iOS, ensuring broad accessibility and usability across diverse user groups, making it a comprehensive platform for both monitoring and intervention in

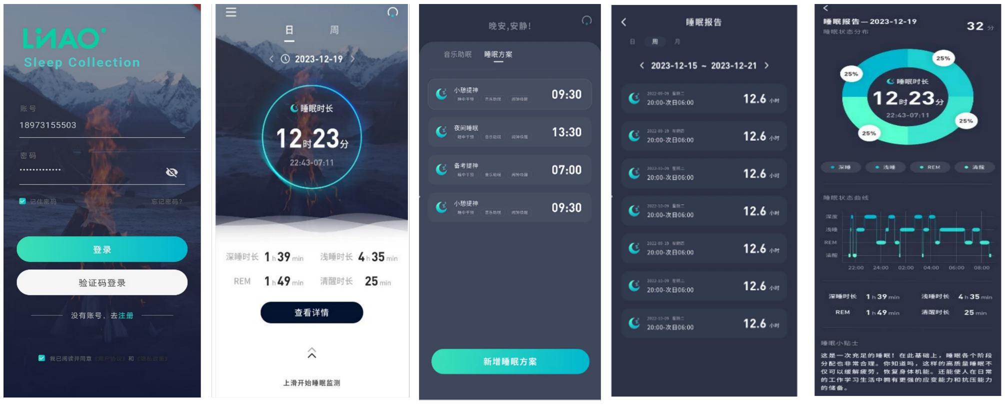


Figure S3: Sleep App interface. The application is available on both Android and iOS platforms.

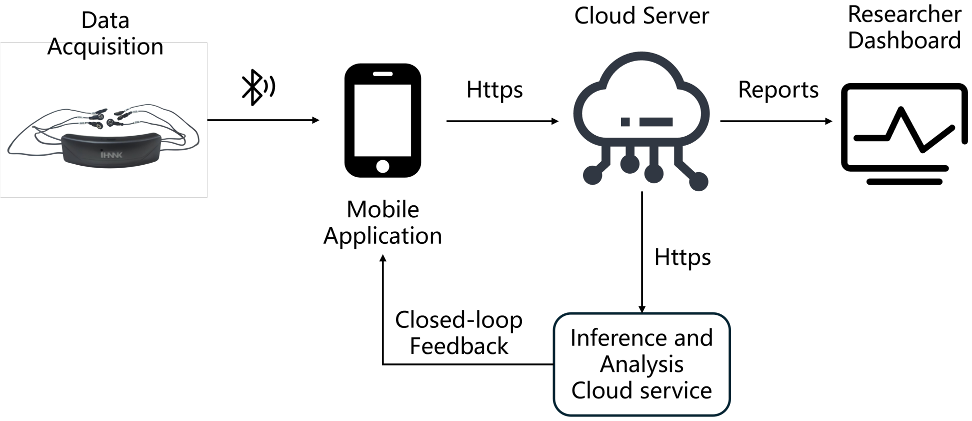


Figure S4: Cloud platform architecture. EEG signals are first acquired via TH25 and transmitted to a cloud processing module. The cloud infrastructure performs real-time data preprocessing, sleep staging, and detection of Sleep events (e.g., spindles, slow waves). Processed results are visualized and accessed by researchers or clinicians through a web-based dashboard or mobile interface.

home-based sleep research.

### Supplementary 3. Cloud platform architecture

As shown in Fig S4, TH25 record real-time EEG , blood oxygen and respiration signals save the data in BDF format. The collection device automatically uploads data to the cloud server through Sleep App(refer Supplementary 3), and the data transmission process is encrypted using HTTPS protocol to ensure privacy and data security.

Based on Python and MNE libraries, automatically load BDF format files and perform filtering and artifact removal processing.Using a Ut-Sleep model(refer Supplementary 4) to achieve automatic sleep staging and detect characteristic waveforms such as spindle waves and slow waves.

### Supplementary 4. Sleep staging and event detection algorithm

The sleep staging and event detection processes in this study were powered by UT-SleepNet, a unified transformer-based semantic segmentation model tailored for high-resolution sleep EEG analysis, with the model structure illustrated in Figure S5. UT-SleepNet is capable of simultaneously performing full-night sleep staging and event-level detection of key microstructural features, including sleep spindles and slow waves.

Unlike traditional classification-based approaches, UT-SleepNet formulates sleep analysis as a dense, time-resolved segmentation task. The model takes preprocessed EEG signals as input and outputs frame-wise labels for both sleep stages (Wake, N1, N2, N3, REM) and waveform events (e.g., spindle onset/offset, slow-wave boundaries).

To ensure generalizability and robustness, UT-SleepNet was trained and evaluated on a large-scale cohort of 9,621 subjects aggregated from six publicly available datasets (SHHS, MROS, SOF, CFS, MESA, and HomePAP). As summarized in Table 2, the model achieved consistent performance in sleep staging across datasets, with macro-averaged accuracy (M-Acc) up to 0.88 and macro-averaged F1-scores (M-F1) up to 0.82. In addition, as shown in Table 3, UT-SleepNet demonstrated high reliability in microstructural event detection, achieving macro-averaged F1-scores of 0.90–0.96 for spindles and 0.91–0.94 for slow waves when evaluated under the IOU = 0.2 criterion (see Figure S6 for the IOU calculation). These results highlight the robustness of the proposed framework and its scalability for large-scale, home-based sleep research.

Table 2: Performance of the proposed model across six public datasets in sleep staging.

| Dataset | Records | Wake | N1 | N2 | N3 | REM | M-Acc | M-F1 |
|---|---|---|---|---|---|---|---|---|
| SHHS | 815 | 0.86 | 0.45 | 0.83 | 0.89 | 0.86 | 0.88 | 0.80 |
| MROS | 324 | 0.82 | 0.45 | 0.79 | 0.85 | 0.82 | 0.87 | 0.76 |
| CFS | 73 | 0.83 | 0.45 | 0.80 | 0.86 | 0.83 | 0.85 | 0.77 |
| HomePAP | 52 | 0.84 | 0.45 | 0.81 | 0.87 | 0.84 | 0.80 | 0.78 |
| MESA | 337 | 0.87 | 0.45 | 0.84 | 0.90 | 0.87 | 0.86 | 0.81 |
| SOF | 112 | 0.88 | 0.45 | 0.85 | 0.91 | 0.88 | 0.87 | 0.82 |

Values represent macro-averaged F1-scores for each sleep stage (Wake, N1, N2, N3, REM), together with macro-averaged accuracy (M-Acc) and macro-averaged F1-score (M-F1). Records indicates the number of subjects included in each dataset.

Table 3: Spindle and slow-wave detection performance across six public datasets.

| **Dataset** | **Spindle Count** | **Slow-wave Count** | **Spindle m-F1** | **Slow-wave m-F1** |
|---|---|---|---|---|
| SHHS | 5724 | 14457 | 0.96 | 0.94 |
| MROS | 2754 | 9463 | 0.92 | 0.93 |
| CFS | 1749 | 6753 | 0.94 | 0.94 |
| HomePAP | 1256 | 4437 | 0.90 | 0.91 |
| MESA | 1946 | 5379 | 0.93 | 0.92 |
| SOF | 729 | 2368 | 0.91 | 0.93 |

Spindle Count and Slow-wave Count represent the total number of detected events across subjects in each dataset. Spindle m-F1 and Slow-wave m-F1 denote the macro-averaged F1-scores of event detection, computed using PSG annotations as the reference standard. True positives (TP) were defined as detected events overlapping with PSG annotations with an intersection-over-union (IOU) greater than 0.2.

### Supplementary 5. Auditory closed-loop stimulation experiment

A total of 19 participants completed the auditory closed-loop stimulation experiment, each undergoing overnight EEG monitoring using the portable TH25 system. Sleep data were recorded from the frontal-mastoid montage (F3–M2), and pink noise stimulation was delivered via a bedside mobile speaker placed approximately 50 cm from the participant's head.

The stimulation protocol followed a real-time closed-loop framework (see Fig. S7 and Fig. S8). EEG signals were continuously transmitted to a cloud-based server, where automatic sleep staging was performed in real time. Upon detection of stable N3 (slow-wave)

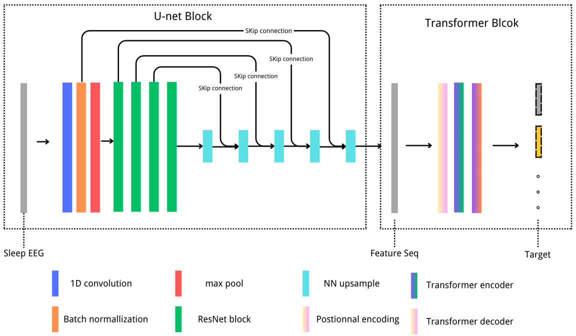


Figure S5: Overall architecture of the proposed **UT-SleepNet**. The model consists of two main components: (i) a U-net block that extracts multi-scale temporal features from raw sleep EEG using convolution, batch normalization, ResNet blocks, max pooling, and nearest-neighbor upsampling with skip connections; and (ii) a Transformer block that models long-range dependencies via positional encoding, Transformer encoder, and Transformer decoder. The final output corresponds to the target sleep stage sequence.

sleep, the stage classification results were returned to the mobile application. Locally, the mobile device applied a 0.5–1.5 Hz bandpass filter to the EEG signal and detected the negative peak (trough) of endogenous slow oscillations. When a trough was identified, a pink noise stimulus (50 ms duration) was triggered after a fixed delay to align with the rising phase of the slow wave.

As shown in Fig. S8, stimulations were only delivered when both conditions were met: (1) the participant was confirmed to be in N3 sleep; and (2) a valid slow-wave trough was detected. To ensure phase-specificity and prevent overstimulation, pink noise was delivered in stimulus pairs with a fixed inter-stimulus interval of 1.075 seconds, consistent with previous studies targeting endogenous slow oscillation frequency. All stimulation events were timestamped and logged for subsequent ERP and behavioral analysis.

Importantly, event-related potential (ERP) and time–frequency analyses revealed that auditory closed-loop stimulation significantly enhanced slow-wave activity (SWA, 0.5–1.5 Hz power spectral density) in the frontal channel. As illustrated in Fig. S9, the SWA power spectrum exhibited a mean increase of **23.7%** in the stimulation condition compared to sham, confirming the efficacy of the TH25-based closed-loop system in boosting endogenous slow oscillations.

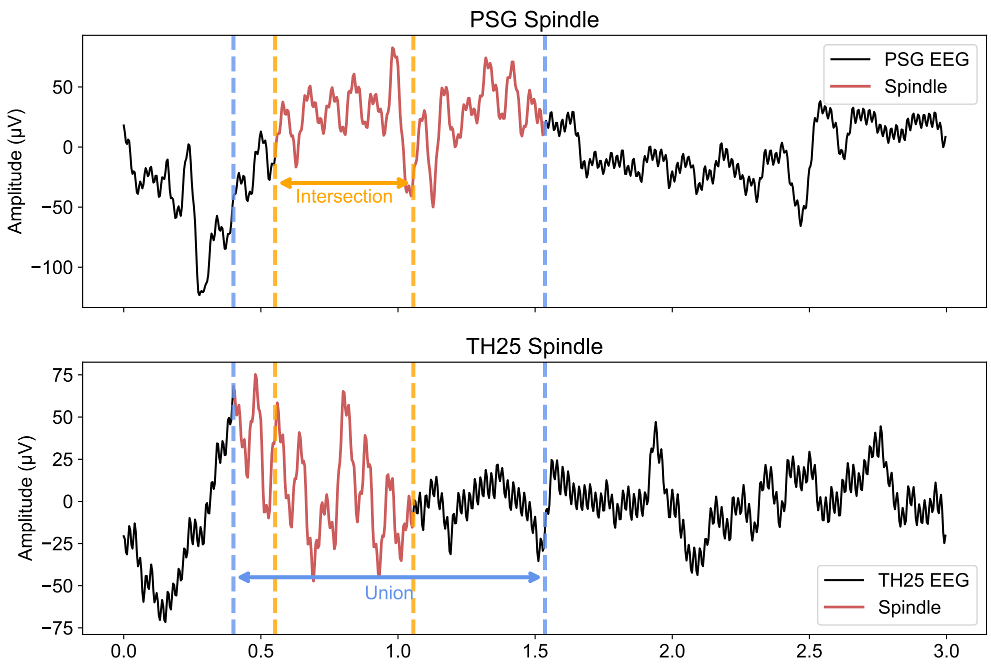


Figure S6: Illustration of spindle event alignment between PSG and TH25 using intersection-over-union (IoU) matching. The top panel shows a PSG-labeled spindle overlaid on its EEG segment, while the bottom panel shows a TH25-labeled spindle with corresponding EEG. The shaded region in each panel denotes the event duration, with the temporal overlap (intersection) and full union used to compute the IoU. A TH25 event is considered a true positive if IoU $> 0.2$.

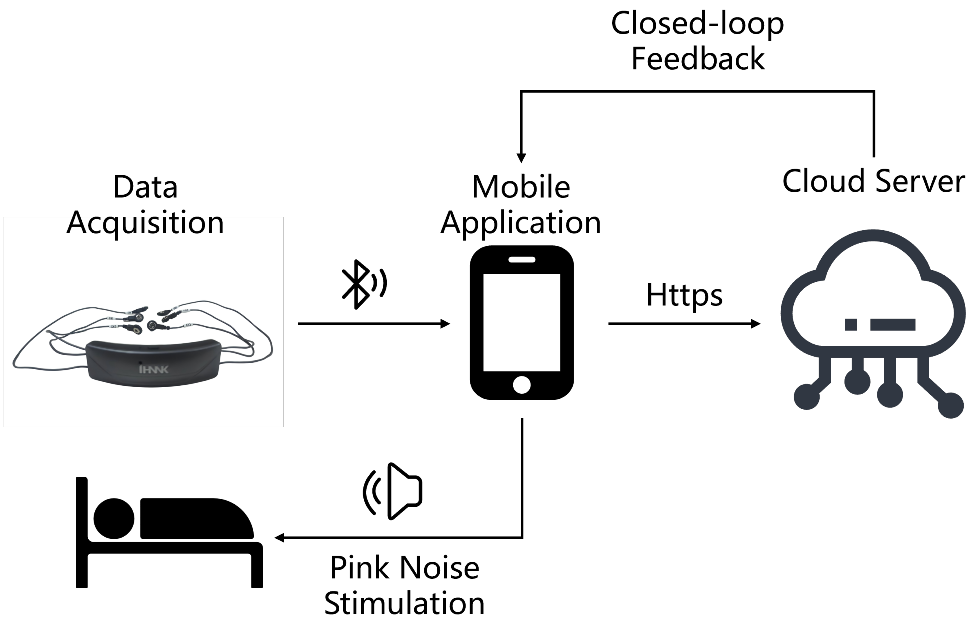


Figure S7: Experimental workflow of the auditory closed-loop stimulation protocol using the TH25 system.

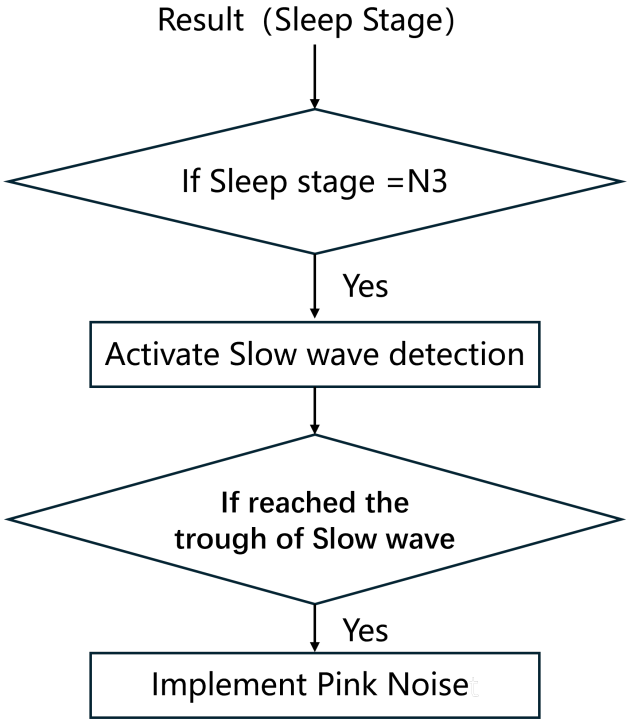


Figure S8: Logical flowchart of pink noise delivery in the auditory closed-loop stimulation protocol. The system continuously monitors EEG in real-time and checks whether the subject is currently in N3 (slow-wave) sleep, based on sleep staging updated every 30 seconds. If in N3, the filtered EEG (0.5–1.5 Hz) is analyzed to detect the negative peak (trough) of endogenous slow oscillations. Upon detecting a trough, the system triggers a pink noise stimulus with a fixed delay to align the onset of stimulation with the up-phase of the slow wave.

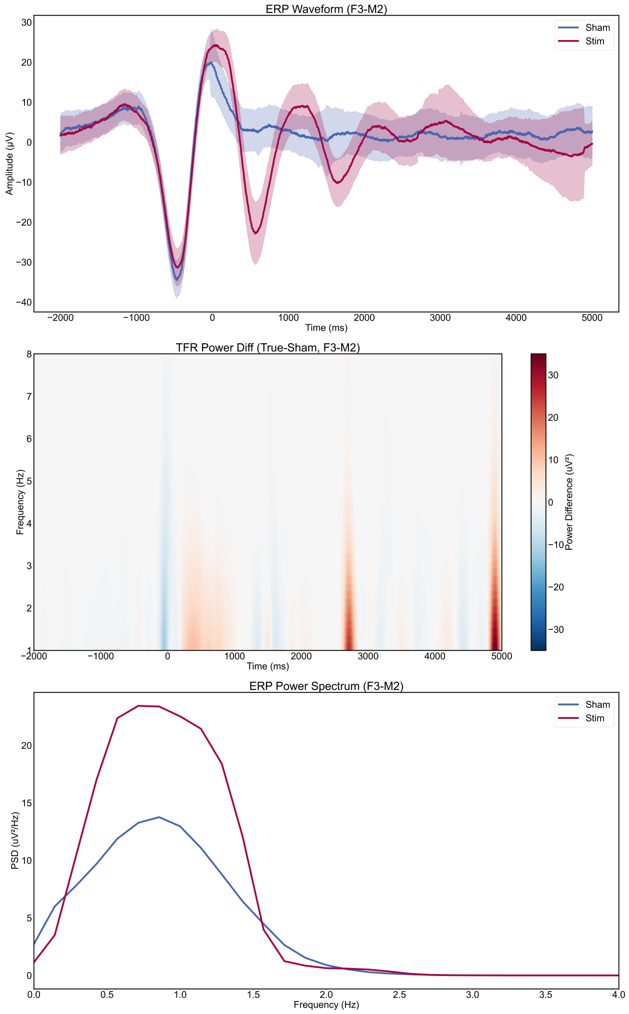


Figure S9: Event-related potential (ERP) responses to closed-loop auditory stimulation during sleep, recorded using the TH25 system at the F3–M2 channel. **(Top)** Averaged ERP waveforms for the stimulation (Stim, red) and sham (blue) conditions, with shaded areas indicating the standard error across trials. The Stim condition shows enhanced slow oscillatory responses, particularly around 0–1200 ms. **(Middle)** Time–frequency representation (TFR) of the power difference between Stim and Sham conditions. Red regions indicate increased low-frequency power under stimulation, especially in the 1–4 Hz (delta) range at multiple time points post-stimulus. **(Bottom)** ERP power spectra show that the Stim condition elicited stronger power in the slow oscillation band (¡1.5 Hz) compared to Sham.
